# Prevalence and Determinants of the Double Burden of Malnutrition among Adolescents: A Cross-Sectional Study in Urban Schools of Nepal

**DOI:** 10.64898/2026.11.02.26367141

**Authors:** Dev Ram Sunuwar, Asmita Shrestha, Ashmita Kiwanayo, Subekshya Karki, Melisha Shrestha, Sudip Sunuwar, Devendra Raj Singh, Kshitij Karki, Narendra Kumar Chaudhary, K.C. Dirghayu

## Abstract

**Background:** The double burden of malnutrition (DBM), characterised by coexisting undernutrition and overnutrition, remains a public health concern among adolescents especially in low- and middle-income countries. Adolescents in Nepal are nutritionally vulnerable, yet evidence on the coexistence of stunting and overweight/obesity is limited. This study assessed the prevalence and associated factors of DBM among school-going adolescents in urban Kathmandu Valley, Nepal.

**Methods:** A school-based cross-sectional study was conducted to assess factors associated with the double burden of malnutrition among adolescents in Kathmandu Valley, Nepal from 25^th^ April, 2022 up to 25^th^ July, 2025. A multi-stage cluster random sampling technique, using the Probability Proportional to Size method, was employed to ensure the selection of the desired number of participants. Students from grades 8 to 10 were randomly selected for face-to-face interviews and anthropometric measurements (height and weight were collected to calculate Body Mass Index (BMI)-for-age Z-scores (BAZ)), using tools adapted to the local context. Bivariate and multivariable models examined the associations between the dependent and independent variables.

**Results:** This study revealed that 15.3% of the students were stunted, 5.5% were underweight, 16.9% were overweight/obese, and 3% of students with the coexistence of DBM.

In the fully adjusted model, the odds of stunting were significantly higher among adolescents aged 15–19 years (aOR = 2.03, 95% CI: 1.35–3.06), salty-snack consumers (aOR = 1.53, 95% CI: 1.00–2.36), those with low nutritional knowledge (aOR = 3.17, 95% CI: 2.17–4.66), and low diet quality (aOR = 7.27, 95% CI: 4.96–10.65). Conversely, fatty-food consumption was associated with lower odds of stunting (aOR = 0.65, 95% CI: 0.42–1.00). For overweight/obesity, older adolescents had significantly lower odds (aOR = 0.49, 95% CI: 0.35–0.67). Regarding the double burden of malnutrition (DBM), low nutritional knowledge (aOR = 2.16, 95% CI: 0.99–4.71) and low diet quality (aOR = 5.27, 95% CI: 2.42–11.48) were significantly associated with higher odds.

**Conclusion:** The double burden of malnutrition exists among adolescents in urban Kathmandu Valley, highlighting a critical public health challenge. Socio-demographic, dietary, and behavioral factors significantly influence its prevalence. Comprehensive school- and community-based interventions are crucial for addressing this dual nutritional burden.

## Introduction

Over the past three decades, malnutrition and its various forms have emerged as a significant challenge to the growing population drawing attention from the global health community^1^. The double burden of malnutrition (DBM) refers to the coexistence of undernutrition (such as stunting, underweight) and overnutrition (overweight or obesity) within the same individual, household, or population level, which is common in low and middle-income countries (LMICs). In adolescents, this often reflects the combined impact of poor dietary quality, rapid nutrition transition, epigenetics, and lifestyle changes during a critical period of growth and development^2 3^.

Globally, an estimated 390 million children and adolescents aged 5–19 years are overweight, including 160 million living with obesity, while approximately 190 million experience underweight, defined as a BMI-for-age more than two standard deviations below the reference median. In 2022, around 149 million children under 5 years were stunted, and 37 million were overweight or obese. Nearly half of all deaths among children under 5 are attributed to undernutrition, with the majority occurring in low- and middle-income countries^3^. The prevalence of overweight and obesity among children and adolescents, aged 5-19 years has increased rapidly from 4% in 1975 to 18% in 2016^4^. Adding to this, a systematic review revealed the prevalence of greater than 14% for overweight/obesity in adolescents^5^. One in three persons residing in Low- and middle-income countries (LMICs) is affected by the DBM^6 7^, resulting in the global nutrition transition from childhood up to adolescence^8^. A study conducted in 57 LMICs reported that 10.2% of adolescents were stunted, 5.5% underweight, 21.4% overweight/obese^1^. By the 2010s, approximately 38 % of the 126 LMICs experienced DBM with a notably high prevalence in sub-Saharan Africa, South Asia, East Asia, and the Pacific^5^. Further, the evidence revealed that 2% of school going adolescents in South Asian countries showed the prevalence of concurrent stunting and overnutrition^9^.

The DBM was mostly prevalent among mother-child pairs, children under five, and women of reproductive age ^10^. Additionally, the prevalence of DBM among mother-child pairs was 6.6% in Nepal^11^ affecting overall health system, future economic growth, and human well-being^8 12^. With around a quarter of the population (24%) are adolescents in Nepal^13^,the Global School-Based Student Health Survey (GSHS) Nepal 2015 report shows that 6.7% of adolescent students were overweight, while 0.6% were obese^12^. Adding to this, the Nepal National Micronutrient Survey (NMSS), 2016, showed that 32.0% of the boys and 32.1% of the girls among adolescents were stunted^14^. The prevalence of overweight ranged from 4-5% among both boys and girls, whilst obesity was below one percent among both sexes^15^. Addressing the nutrition needs of adolescents is essential to break the vicious cycle of intergenerational malnutrition, and chronic diseases^16^.

Different anthropometric indicators provide insight into nutritional deficiencies. Stunting is widely recognized as a marker of chronic undernutrition, reflecting long-term health consequences, while obesity indicates excess nutrition and an increased risk of non-communicable diseases such as type 2 diabetes, hypertension, and cardiovascular disease^17 18^. Wasting is more indicative of acute malnutrition driven by recent dietary intake and illness^3^. Adolescent’s higher nutritional needs, their rapid physical and physiological development puts them at a higher risk of malnutrition ^11^. Additionally, the DBM poses several long-term complications, especially if developed during childhood and adolescence, as it increases the risk of diverse chronic non-communicable diseases (NCDs), whose development is closely associated with dietary habits and lifestyle factors^5^. With a lower mortality rate than any other age group and a relatively low morbidity rate, adolescents are generally not a priority for public health interventions^10^. By 2030, an estimated 150 million individuals aged 10–19 years worldwide are expected to be living with obesity^19^. In children and adolescents, obesity is a chronic health condition arising from a multifaceted interaction of physiological, psychological, environmental, socioeconomic, and genetic influences^20^.

The growing burden of DBM among adolescents is largely attributed to the nutrition transition-shift in dietary habits and lifestyles driven by urbanization and economic development. It includes increased consumption of ultra-processed foods high in sugar, salt, trans-fats, and calories, alongside decreased intake of nutrient dense foods and reduced physical activity. It has led to shift in nutritional outcomes from undernutrition to overnutrition and now double burden of under and over nutrition ^11 18^. These ultra-processed foods are likely to be cheaper and easily available but of poor nutritional quality, exacerbating both under and over nutrition risks of malnutrition^18^. Rapid economic transformation, urbanization, cultural expectations for body size and sedentary behavior are some of the key drivers of DBM in this population^21^. Government of Nepal has implemented various plans, policies, and programs to reduce adolescent malnutrition like Multi-Sector Nutrition Plan II^22^, and school health nutrition program^23^. However, many efforts often prioritize focus on undernutrition among children under five and women of reproductive age, leaving adolescents relatively underserved. There are also implementation challenges, as the limited cross sector collaboration, insufficient data on adolescent malnutrition, particularly regarding DBM.

Despite the growing burden and significant long-term health risks of the DBM, research among adolescents in Nepal remains limited. Existing research primarily addresses household or population level DBM or focuses on younger children and mother^24 25^. To address this gap, this study aims to assess the prevalence and determinants of DBM-defined as concurrent stunting and overweight - among adolescents in urban Kathmandu Valley, contributing to provide evidence needed to improve the adolescent nutrition policy and programing in Nepal.

## Materials and Methods

### Study design, sites, and participants

A school-based cross-sectional study design was used to assess the factors associated with DBM among adolescents in Kathmandu Valley, Nepal. Kathmandu Valley is the central lower part standing at 1,425 meters (4,675 ft) above sea level. It consists of three districts (Kathmandu, Lalitpur, and Bhaktapur) of Bagmati Province and is the most developed as well as the largest urban agglomeration of Nepal^26^ with a total population of 6, 116,866^27^ and a total area of 933.73 square kilometers (360.52 sq mi), with an annual population growth rate of 0.97%^27^.

The study was conducted in schools of Kathmandu, Lalitpur, and Bhaktapur districts from 25^th^ April, 2022 up to 25^th^ July, 2025. There were 1,362 schools in Kathmandu, 475 in Lalitpur, and 344 in Bhaktapur. Among the total schools, 616 schools were community schools, and the remaining 1,565 schools were institutional ^28^. Adolescent students, both male and female, aged 10-19 years studying in the selected schools of Kathmandu Valley were the study population for this study. Details on study sites have been presented on the map (Fig.1).

**Figure 1.**
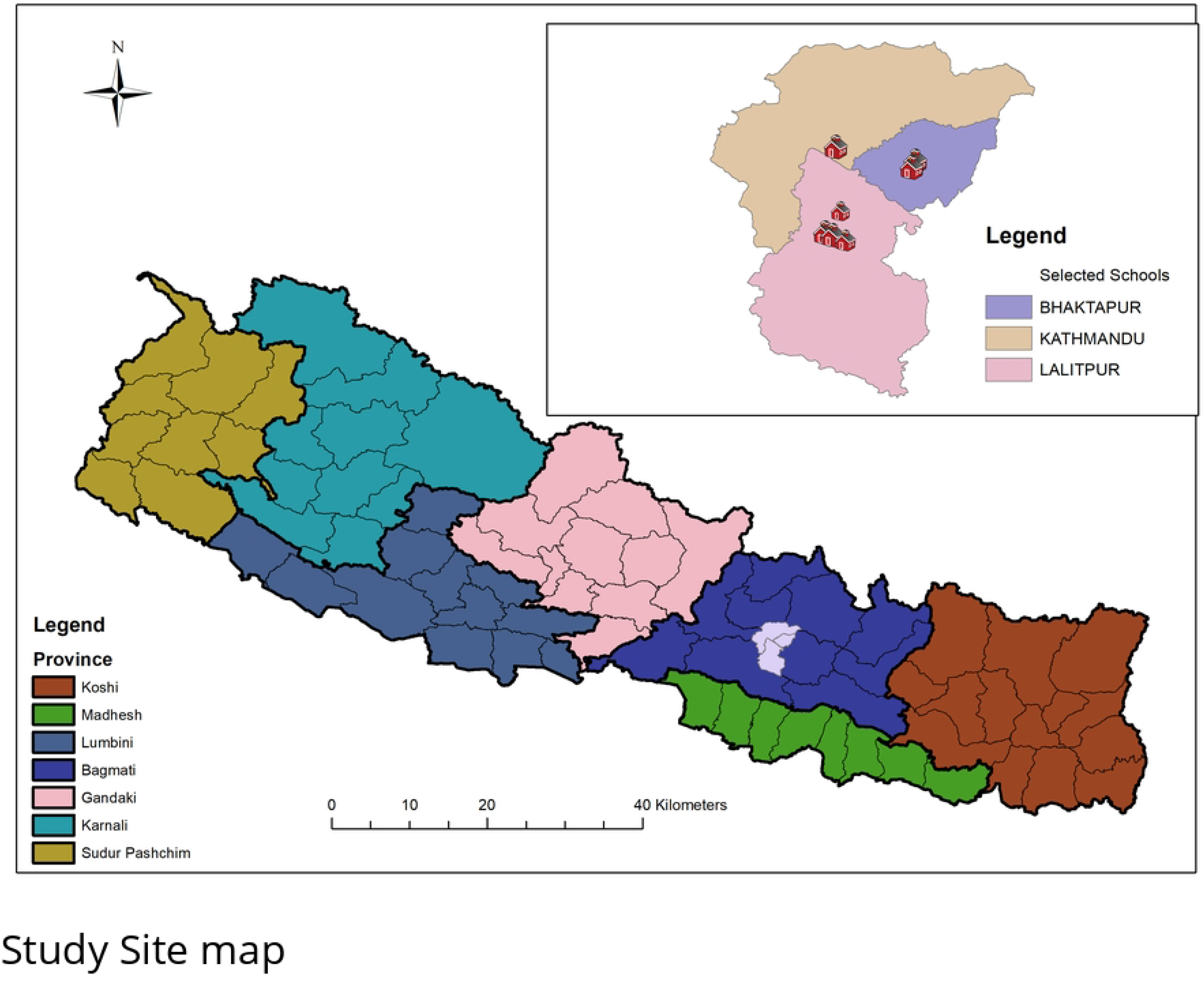
Map showing the locations of selected schools in Kathmandu Valley, Nepal.

The map was created using ArcGIS Pro (version 3.4). The base files of the administrative provinces and districts of Nepal were obtained from the Government of Nepal, Ministry of Land Management, and Natural Earth, which are publicly available without restrictions (http://dos.gov.np/nepal-map).

Although most students in grades 8–10 typically fall between the ages of 12 and 17, the study included all enrolled students aged 10–19 to align with the WHO definition of adolescence and to capture any students who may have started school earlier or experienced grade repetition. This broader age range allowed for a comprehensive assessment of nutritional status across the full spectrum of adolescence. Both males and females aged 10– 19 years studying grade 8–10 class and those who gave consent to participate in the study were included in the study, whereas adolescents who cannot be traced during the data collection stage and who are severely ill at the time of data collection were excluded.

### Sample size and sampling technique

The sample was determined using a single proportional formula with a conservative prevalence of 0.5 considering key indicators. Taking the standard normal deviation (Zα) at 95% confidence level, and 4% margin of error (d), the minimum sample size was calculated using the formula: Sample size (n0) = Z2 ×p (1-p)/d2, where Z = 1.96 (for 95% confidence), p = 0.5 (conservative estimate), and d = 0.04. This yielded a calculated sample size (n₀) of 600. After adjusting for a 10% non-response rate and a design effect of 2, the final total sample size was 1,320. A multi-stage random sampling technique was used to select participants for this study. First, three wards from each district (Kathmandu, Lalitpur, Bhaktapur) were randomly selected. Next, all secondary schools (grades 8–10) within these wards were listed, and eight schools were chosen to ensure representation from both public and private institutions. Specifically, one public school was selected from Kathmandu, two public and two private schools from Lalitpur, and two public and one private school from Bhaktapur. In the third stage, students from grades 8–10 in these schools were selected using Probability Proportional to Size (PPS) sampling. Finally, simple random sampling was employed to select students from the list of eligible adolescents in the chosen schools.

### Data collection tools and techniques

#### Data collection tool

**Semi-structured Questionnaire**: The socio-demographic information was collected using a semi-structured questionnaire adapted from the global school-based student health survey (GSHS) questionnaire^12^.

**Behavior factors:** The questionnaire on smoking, alcohol, drug abuse, and sedentary behavior was also adapted from the GSHS questionnaires^12^. These questions asked if the participants had consumed any alcohol, drugs, or smoking in the last 30 days and the frequency of consumption^12^.

**Physical activity:** The physical activity questionnaire was adapted from the Nepalese version of the GSHS questionnaire mode^12^.

**Dietary Habit**: Dietary habits were assessed by adopting GSHS questionnaire modules developed by the WHO^12^. The information was collected to assess the dietary habits of adolescents on consumption of different types of foods (fruits consumption per day, vegetables consumption, food with high fats), frequency of junk food consumption, carbonated soft drinks (Coke, Pepsi, Sprite), fast food (Momo; a popular Nepali dumpling, Pizza; a traditional Nepali recipe made of rice flour etc.) consumed in last 30 days from the date of data collection.

**Nutritional knowledge:** Nutritional knowledge level was determined based on the eighteen questions (Q1-Q18). A nutrition questionnaire developed in Polish conditions and education based on the Polish Food Guide Pyramid was adapted and contextualised into Nepalese terms^29^. The score ‘1’ was given for correct answers, and a ‘0’ was given for wrong or unknown answers and missing data. Respondents were divided into three nutritional knowledge levels: lowest (0-4 points), moderately low (5-7 points), and higher (8-10 points). This tool has already been utilized and implemented previously in Nepal^30^.

**Diet quality**: Diet quality was assessed through 11 food-frequency questions focused on both healthy (e.g., fruits, dairy, vegetables) and unhealthy (e.g., soft drinks, fast food) items. Two indices were generated: the pro-Healthy Diet Index (pHDI) and non-Healthy Diet Index (nHDI), based on consumption frequency. Final diet quality was categorised as low, moderate, or high following established scoring cut-offs on the basis of usual food frequency consumption within the last 12 months ^31^ and has already been implemented in previous research in Nepal with similar settings^30^.

**Anthropometric measurement:** Height was measured using a stadiometer to the nearest 1cm, whereas participants’ weight was measured using the Seca weighing scale to the nearest 0.1kg.

### Data collection techniques

The questionnaire was developed, translated into Nepali, and validated by language experts. Back translation was done before pre-testing, which involved 10% of the study sample and was subsequently adjusted. Face-to-face interviews and anthropometric measurements were conducted using adapted tools in the local context. Research assistants were trained in data collection, sampling, and ethical guidelines. Piloting ensured data consistency, with field supervisors overseeing the process. Semi-structured questionnaires gathered socio-demographic, behavioral, and nutritional data. Apart from Face-to-face interviews, an anthropometric and nutrition survey was conducted to assess the adolescents’ nutritional status utilizing the height-for-age (HAZ) and BMI-for-age (BMIZ) indices as Z-scores^31^. Validity and reliability were ensured using a random sampling process at each stage to ensure representative participants and was maintained throughout data collection, entry, and analysis.

### Study variables

#### Dependent variables

The primary dependent variables were undernutrition (stunting and underweight), overnutrition (overweight and obesity), and the DBM, coexistence of undernutrition (stunting) and overnutrition (overweight/obesity).

Stunting was defined as height-for-age Z score (HAZ) < −2SD, and underweight was defined as BMI-for-age Z score (BAZ) < −2SD^32^. Overweight was defined as BAZ > +1SD and obesity as BAZ > +2SD, following WHO growth reference standards 2006 for children and adolescents aged 5–19 years^33^. Anthropometric z-scores (HAZ and BAZ) were calculated using the WHO AnthroPlus software^34^. Overweight and obesity were grouped together in analysis due to the relatively low prevalence of obesity alone, which would have limited statistical power for separate modelling. This approach is commonly used in similar studies examining adolescent nutritional status in LMICs^9 35^. The double burden of malnutrition was operationalized as the concurrent presence of stunting (HAZ < −2SD) and overweight/obesity (BAZ > +1SD)^36^. For analysis, this co-occurrence was analyzed as a binary outcome, coded as “Yes” if both stunting and overweight/obesity were present and “No” if this combination was absent. This approach, adapted from prior study^37^, allowed us to clearly distinguish adolescents experiencing DBM from those with only one or neither form of malnutrition.

#### Independent variables

The independent variables were categorized into three domains: socio-demographic factors (age, sex, grade, caste, religion, family type, parents’ education level, and parents’ occupation), behavioral factors (physical activity, number of physical education class days, and sedentary behavior), and dietary patterns (intake of seasonal fruits and vegetables, carbonated drinks, fast food, salty food, and fatty food, along with nutritional knowledge and overall diet quality). These variables were included based on their established associations with adolescent nutritional status in previous literature and their biological plausibility in influencing stunting, underweight, overweight/obesity, and the double burden of malnutrition^38 39^. Variables were categorized, and in some cases merged, to ensure meaningful interpretation and address sparse data in individual categories, thereby improving the robustness of statistical analysis.

### Data management and analysis

The collected data were entered into EpiData version 3.0 by four trained data entry professionals and data was analyzed using Stata/MP version 14.1 (StataCorp LP, College Station, Texas). Frequency, and percentage were calculated using descriptive statistics. Although BMI-for-age and height-for-age z-scores were collected as continuous variables, we categorized them using WHO cut-offs to define underweight, stunting, overweight, and obesity. The co-occurrence was treated as a binary outcome (“Yes” if both stunting and overweight/obesity were present, “No” otherwise) to ensure clear classification and comparability with previous literatures^40 41^. Using a binary outcome also facilitates binary logistic regression analysis, allowing for the estimation of odds ratios for factors associated with DBM, and provides a straightforward interpretation for public health planning and policy. Multivariable logistic regressions assessed the association between predictors and outcome variables. Three adjusted models were constructed to evaluate the associations. Model 1 was adjusted for demographic factors, Model 2 for behavioral and dietary factors, and Model 3 for demographic, behavioral, and dietary factors. All behavioral and dietary pattern variables were included regardless of their bivariate significance, as they are biologically and theoretically important contributors to adolescent nutritional status and help control for potential confounding. Multicollinearity was assessed using variance inflation factors (VIF) to prevent statistical bias, and model fit was evaluated using the Hosmer-Lemeshow goodness-of-fit test. The mean value of VIF < 10 was the cut-off point^43^. Crude odds ratio (cOR) and adjusted odds ratio (aOR) with 95% CIs were calculated to assess the presence and strength of association, with a p < 0.05 used for the determination of statistical significance.

#### Ethical aspects

Approval for this study was sought from the Nepal Health Research Council (Protocol registration number: 144\2022p). In addition, we obtained formal written permission letters from each participating school to conduct the study. Before data collection, permission was obtained from schools, written informed consent from guardians, and written assent from participants. The study’s purpose was clearly explained to all. No personal identifying information was collected, participation was voluntary, and confidentiality was strictly maintained throughout the study.

### Patient and Public involvement

Patient and public weren’t involved in design, implementation, reporting and dissemination plans of this research.

## Results

The findings revealed that 15.3% of the students were stunted, 5.5% were underweight, and 16.9% were overweight/obese. Additionally, the coexistence of DBM was observed in 3.0% of the students **(Table 1).** In terms of socio-demographic characteristics, male adolescents slightly outnumbered females (50.3% vs. 49.7%). More than half (54.9%) were aged 15–19 years, and the majority were from disadvantaged ethnic groups (60.5%). The majority (86.8%) identified as Hindu.

**Table 1.** Socio demographic characteristics, behavioral factors, and dietary pattern of participants.

| **Variables** | **Frequency (N=1324)** | **Percentage** | **Stunting** |  | **DBM** |  |
|---|---|---|---|---|---|---|
|  | **(n)** | **(%)** | **Not stunted**<br>**n (%)** | **Yes**<br>**n (%)** | **Not DBM**<br>**n (%)** | **Yes**<br>**n (%)** |
| **Socio demographic factors** |  |  |  |  |  |  |
| **Participants sex** |  |  |  |  |  |  |
| Male | 666 | 50.30 | 567 (85.14) | 99 (14.86) | 600 (97.09) | 18 (2.91) |
| Female | 658 | 49.70 | 554 (84.19) | 104 (15.81) | 613 (96.84) | 20 (3.16) |
| **Age category** |  |  |  |  |  |  |
| 10-14 | 596 | 45.02 | 531 (89.09) | 65 (10.91) | 556 (96.70) | 19 (3.30) |
| 15-19 | 728 | 54.98 | 590 (81.04) | 138 (18.96) | 657 (97.19) | 19 (2.81) |
| **Grade** |  |  |  |  |  |  |
| Class 8 | 259 | 19.56 | 210 (81.08) | 49 (18.92) | 238 (95.58) | 11 (4.42) |
| Class 9 | 789 | 59.59 | 684 (86.69) | 105 (13.31) | 724 (97.31) | 20 (2.69) |
| Class 10 | 276 | 20.85 | 227 (82.25) | 49 (17.75) | 251 (97.29) | 7 (2.71) |
| **Caste** |  |  |  |  |  |  |
| Disadvantaged ethnic group | 802 | 60.57 | 667 (83.17) | 135 (16.83) | 741 (96.36) | 28 (3.64) |
| Advantaged ethnic group | 522 | 39.43 | 454 (86.97) | 68 (13.03) | 472 (97.93) | 10 (2.07) |
| **Religion** |  |  |  |  |  |  |
| Hindu | 1150 | 86.86 | 984 (85.57) | 166 (14.43) | 1057 (97.24) | 30 (2.76) |
| Non-Hindu | 174 | 13.14 | 137 (78.74) | 37 (21.26) | 156 (95.12) | 8 (4.88) |
| **Family Type** |  |  |  |  |  |  |
| Nuclear | 597 | 45.09 | 522 (87.44) | 75 (12.56) | 548 (97.86) | 12 (2.14) |
| Extended | 727 | 54.91 | 599 (82.39) | 128 (17.61) | 665 (96.24) | 26 (3.76) |
| **Mother education** |  |  |  |  |  |  |
| Low | 447 | 37.88 | 366(81.88) | 81(18.12) | 408(95.55) | 19(4.45) |
| High | 733 | 62.12 | 636(86.77) | 97(13.23) | 671(98.10) | 13(1.90) |
| **Father education** |  |  |  |  |  |  |
| Low | 281 | 23.73 | 223(79.36) | 58 (20.64) | 258(94.16) | 16(5.84) |
| High | 903 | 76.27 | 782(86.60) | 121 (13.40) | 822(97.74) | 19(2.26) |
| **Father occupation** |  |  |  |  |  |  |
| Job/Business | 967 | 73.04 | 836(86.45) | 131(13.55) | 894(97.70) | 21(2.30) |
| Agriculture | 357 | 26.96 | 285(79.83) | 72(20.17) | 319(94.94) | 17(5.06) |
| **Mother occupation** |  |  |  |  |  |  |
| Job/Business | 546 | 41.24 | 478(87.55) | 68(12.45) | 507(97.69) | 12(2.31) |
| Agriculture | 778 | 58.76 | 643(82.65) | 135(17.35) | 706(96.45) | 26(3.55) |
| **Behavioral factors** |  |  |  |  |  |  |
| **Physical Activity** |  |  |  |  |  |  |
| Not Active | 400 | 30.21 | 329 (82.25) | 71 (17.75) | 362 (96.28) | 14 (3.72) |
| Active | 924 | 69.79 | 792 (85.71) | 132 (14.29) | 851 (97.26) | 24 (2.74) |
| **Ride to or from school** |  |  |  |  |  |  |
| Walking | 943 | 71.22 | 786 (83.35) | 157 (16.65) | 861 (96.63) | 30 (3.37) |
| Vehicle | 381 | 28.78 | 335 (87.93) | 46 (12.07) | 352 (97.78) | 8 (2.22) |
| **Attended physical education class** |  |  |  |  |  |  |
| None | 851 | 64.27 | 721(84.72) | 130(15.28) | 775(96.75) | 26(3.25) |
| Yes | 473 | 35.73 | 400(84.57) | 73(15.43) | 438(97.33) | 12(2.67) |
| **Sedentary behavior** |  |  |  |  |  |  |
| <1hr | 239 | 18.10 | 201 (84.10) | 38 (15.90) | 224 (96.55) | 8 (3.45) |
| 1-4hr | 948 | 71.60 | 805 (84.92) | 143 (15.08) | 859 (96.95) | 27 (3.05) |
| >=5hr | 137 | 10.30 | 115 (83.94) | 22 (16.06) | 130 (97.74) | 3 (2.26) |
| **Dietary pattern** |  |  |  |  |  |  |
| **Seasonal fruits intake** |  |  |  |  |  |  |
| Never | 471 | 35.57 | 379 (80.47) | 92 (19.53) | 429(95.76) | 19(4.24) |
| Yes | 853 | 64.43 | 742 (86.99) | 111 (13.01) | 784(97.63) | 19(2.37) |
| **Seasonal Vegetables intake** |  |  |  |  |  |  |
| Never | 274 | 20.69 | 220(80.29) | 54(19.71) | 251(94.72) | 14(5.28) |
| Yes | 1050 | 79.31 | 901(85.81) | 149(14.19) | 962(97.57) | 24(2.43) |
| **Carbonated drinks intake** |  |  |  |  |  |  |
| Never | 951 | 71.83 | 807(84.86) | 144(15.14) | 880(97.24) | 25(2.76) |
| Yes | 373 | 28.17 | 314(84.18) | 59(15.82) | 333(96.24) | 13(3.76) |
| **Fast foods intake** |  |  |  |  |  |  |
| Never | 739 | 55.82 | 628(84.98) | 111(15.02) | 682(96.88) | 22(3.13) |
| Yes | 585 | 44.18 | 493(84.27) | 92(15.73) | 531(97.07) | 16(2.93) |
| **Salty foods intake** |  |  |  |  |  |  |
| Never | 587 | 44.34 | 511(87.05) | 76(12.95) | 540(96.60) | 19(3.40) |
| Yes | 737 | 55.66 | 610(82.77) | 127(17.23) | 673(97.25) | 19(2.75) |
| **Fatty foods intake** |  |  |  |  |  |  |
| Never | 733 | 55.36 | 609(83.08) | 124(16.92) | 680(97.14) | 20(2.86) |
| Yes | 591 | 44.64 | 512(86.63) | 79(13.37) | 533(96.73) | 18(3.27) |
| **Hunger due to lack of food** |  |  |  |  |  |  |
| Never | 1000 | 75.53 | 847(84.70) | 153(15.30) | 918(97.14) | 27(2.86) |
| Yes | 324 | 24.47 | 274(84.57) | 50(15.43) | 295(96.41) | 11(3.59) |
| **Nutrition Knowledge** |  |  |  |  |  |  |
| Low | 410 | 30.97 | 292(71.22) | 118(28.78) | 364(94.55) | 21(5.45) |
| High | 914 | 69.03 | 829(90.70) | 85(9.30) | 849(98.04) | 17(1.96) |
| **Diet quality** |  |  |  |  |  |  |
| Low | 278 | 21.00 | 162(58.27) | 116(41.73) | 244(92.08) | 21(7.92) |
| High | 1046 | 79.00 | 959(91.68) | 87(8.32) | 969(98.28) | 17(1.72) |
| **Prevalence** |  |  |  |  |  |  |
| **Stunting** |  |  |  |  |  |  |
| Not stunted | 1121 | 84.67 |  |  |  |  |
| Stunted | 203 | 15.33 |  |  |  |  |
| **BMI four classification** |  |  |  |  |  |  |
| Underweight | 73 | 5.51 |  |  |  |  |
| Normal | 1026 | 77.49 |  |  |  |  |
| Overweight | 170 | 12.84 |  |  |  |  |
| Obese | 55 | 4.15 |  |  |  |  |
| **Coexistence of DBM** |  |  |  |  |  |  |
| Not DBM | 1213 | 96.96 |  |  |  |  |
| DBM | 38 | 3.04 |  |  |  |  |
|  | Mean | SD |  |  |  |  |
| Height-for-Age Z-score (HAZ) | -0.79 | 1.06 |  |  |  |  |
| BMI-for-Age Z-score (BAZ) | -0.09 | 1.18 |  |  |  |  |

Regarding physical activity, 71.6% of adolescents led a sedentary lifestyle, although a similar proportion (71.22%) reported walking to and from school. In terms of dietary habits, more than half consumed seasonal fruits (64.43%) and vegetables (79.31%). A significant majority reported never consuming carbonated drinks (71.83%), while a little over half reported the same for fast food (55.82%) and fatty foods (55.36%). More than two-thirds had high nutritional knowledge (69.03%), and a substantial majority had a high-quality diet (79%), yet almost one-fourth (24.47%) had experienced hunger due to food shortages **(Table 1).**

### Factors associated with stunting

In the fully adjusted model (Model 3), age, salty-snack consumption, fatty-food consumption, nutritional knowledge, and overall diet quality were significantly associated with stunting among school-going adolescents. Adolescents aged 15–19 years had approximately twice the odds of being stunted compared with those aged 10–14 years (AOR = 2.03, 95% CI: 1.35–3.06; p<0.001). Dietary and nutritional characteristics showed particularly strong associations. Adolescents who consumed salty snacks had 53% higher odds of stunting compared with those who did not (AOR = 1.53, 95% CI: 1.00–2.36; p=0.031). In contrast, fatty-food consumption was associated with lower odds of stunting (AOR = 0.65, 95% CI: 0.42– 1.00; p=0.049). Adolescents with low nutritional knowledge had more than three times higher odds of stunting compared with those with high nutritional knowledge (AOR = 3.17, 95% CI: 2.17–4.66; p<0.001). The strongest association was observed for diet quality: adolescents with low diet quality had more than seven times higher odds of stunting than those with high diet quality (AOR = 7.27, 95% CI: 4.96–10.65; p<0.001).

Among demographic characteristics, sex, caste, parental education, and maternal occupation were not significantly associated with stunting in the fully adjusted model. Similarly, physical activity, physical education class attendance, fruit and vegetable consumption, soft-drink consumption, and fast-food consumption were not significantly associated with stunting. Although paternal agricultural occupation was associated with higher odds of stunting in the demographic model (AOR = 1.58, 95% CI: 1.08–2.31; p=0.019), this association was attenuated and became non-significant after adjustment for behavioral and dietary factors (AOR = 1.38, 95% CI: 0.89–2.13; p=0.146) **(Table 2).**

**Table 2.** Multivariable binary logistic regression analyses for factors associated with stunting among school-going adolescents in Kathmandu Valley, Nepal.

| **Predictor** |  | **Model 1** |  | **Model 2** |  | **Model 3** |  |
|---|---|---|---|---|---|---|---|
| Demographic factors | **Category** | **Adjusted OR (95% CI)** | **p-value** | **Adjusted OR (95% CI)** | **p-value** | **Adjusted OR (95% CI)** | **p-value** |
| Sex | Male (Ref) | — | — |  |  | — | — |
|  | Female | 1.16 (0.83, 1.62) | 0.398 |  |  | 1.10 (0.75, 1.61) | 0.621 |
| Age category | 10–14 (Ref) | — | — |  |  | — | — |
|  | 15–19 | 1.93 (1.35, 2.78) | <0.001* |  |  | 2.03 (1.35, 3.06) | <0.001* |
| Caste category | Advantage ethnic group (Ref) | — | — |  |  | — | — |
|  | Disadvantage ethnic group | 1.17 (0.82, 1.68) | 0.384 |  |  | 1.02 (0.68, 1.54) | 0.917 |
| Mother’s education | High (Ref) | — | — |  |  | — | — |
|  | Low | 1.05 (0.72, 1.53) | 0.807 |  |  | 0.91 (0.59, 1.39) | 0.652 |
| Father’s education | High (Ref) | — | — |  |  | — | — |
|  | Low | 1.27 (0.84, 1.93) | 0.254 |  |  | 1.08 (0.67, 1.74) | 0.759 |
| Mother’s occupation | Job/business (Ref) | — | — |  |  | — | — |
|  | Agriculture | 1.07 (0.74, 1.53) | 0.732 |  |  | 1.07 (0.72, 1.61) | 0.733 |
| Father’s occupation | Job/business (Ref) | — | — |  |  | — | — |
|  | Agriculture | 1.58 (1.08, 2.31) | 0.019 |  |  | 1.38 (0.89, 2.13) | 0.146 |
| Behavioral and dietary factors |  |  |  |  |  |  |  |
| Physical activity | Yes (Ref) |  |  | Ref | — | — | — |
|  | No |  |  | 1.02 (0.71, 1.46) | 0.904 | 1.11 (0.74, 1.66) | 0.626 |
| PE class attendance | Yes (Ref) |  |  | — | — | — | — |
|  | No |  |  | 1.16 (0.81, 1.66) | 0.415 | 1.12 (0.74, 1.68) | 0.602 |
| Fruit consumption | Yes (Ref) |  |  | — | — | — | — |
|  | No |  |  | 1.18 (0.81, 1.72) | 0.393 | 1.04 (0.67, 1.62) | 0.86 |
| Vegetable consumption | Yes (Ref) |  |  | — | — | — | — |
|  | No |  |  | 1.09 (0.71, 1.67) | 0.687 | 1.09 (0.68, 1.76) | 0.715 |
| Soft drink consumption | No (Ref) |  |  | — | — | — | — |
|  | Yes |  |  | 1.05 (0.69, 1.60) | 0.832 | 1.12 (0.70, 1.81) | 0.629 |
| Fast food consumption | No (Ref) |  |  | — | — | — | — |
|  | Yes |  |  | 1.01 (0.69, 1.48) | 0.943 | 1.02 (0.67, 1.56) | 0.923 |
| Salty snacks consumption | No (Ref) |  |  | — | — | — | — |
|  | Yes |  |  | 1.62 (1.10, 2.38) | 0.014 | 1.53 (1.00, 2.36) | 0.031* |
| Fatty food consumption | No (Ref) |  |  | — | — | — | — |
|  | Yes |  |  | 0.79 (0.54, 1.15) | 0.221 | 0.65 (0.42, 1.00) | 0.049* |
| Nutritional Knowledge | High (Ref) |  |  | — | — | — | — |
|  | Low |  |  | 3.20 (2.27, 4.50) | <0.001 | 3.17 (2.17, 4.66) | <0.001* |
| Diet quality | High(Ref) |  |  | — | — | — | — |
|  | Low |  |  | 6.88 (4.90, 9.67) | <0.001 | 7.27 (4.96, 10.65) | <0.001* |

*\*Statistically significant at p < 0.05. Model 1 is adjusted for demographic factors; Model 2 is adjusted for behavioral and dietary factors; and Model 3 is adjusted for both demographic and dietary factors.*

### Factors associated with body weight status

In the fully adjusted model, age was the only significant predictor of body weight status. Adolescents aged 15–19 years had significantly lower odds of becoming overweight/obese compared with those aged 10–14 years (AOR = 0.49, 95% CI: 0.35–0.67; p<0.001), indicating approximately 51% lower odds among older adolescents. No significant associations were observed for sex, caste, parental education or occupation, physical activity, physical education attendance, individual dietary behaviors, nutritional knowledge, or overall diet quality.

**Table 3.** Multivariable binary logistic regression analyses for factors associated with body weight status among school-going adolescents in Kathmandu Valley, Nepal.

|  |  | **Model 1** |  | **Model 2** |  | **Model 3** |  |
|---|---|---|---|---|---|---|---|
| **Predictor** | **Category** | **Adjusted OR (95% CI)** | **p-value** | **Adjusted OR (95% CI)** | **p-value** | **Adjusted OR (95% CI)** | **p-value** |
| Sex | Male (Ref) | — | — |  |  | — | — |
|  | Female | 0.90 (0.66, 1.24) | 0.519 |  |  | 0.93 (0.67, 1.29) | 0.663 |
| Age category | 10–14 (Ref) | — | — |  |  | — | — |
|  | 15–19 | 0.47 (0.34, 0.65) | <0.001* |  |  | 0.49 (0.35, 0.67) | <0.001* |
| Caste | Advantage ethnic group (Ref) | — | — |  |  | — | — |
|  | Disadvantage ethnic group | 1.01 (0.73, 1.41) | 0.948 |  |  | 1.00 (0.71, 1.41) | 0.985 |
| Mother's education | High (Ref) | — | — |  |  | — | — |
|  | Low | 1.01 (0.70, 1.47) | 0.962 |  |  | 0.99 (0.68, 1.45) | 0.974 |
| Father's education | High (Ref) | — | — |  |  | — | — |
|  | Low | 1.00 (0.65, 1.54) | 0.983 |  |  | 0.96 (0.62, 1.50) | 0.867 |
| Mother's occupation | Job/business (Ref) | — | — |  |  | — | — |
|  | Agriculture | 0.88 (0.63, 1.22) | 0.433 |  |  | 0.86 (0.61, 1.20) | 0.379 |
| Father's occupation | Job/business (Ref) | — | — |  |  | — | — |
|  | Agriculture | 1.01 (0.68, 1.51) | 0.952 |  |  | 1.00 (0.66, 1.50) | 0.989 |
| **Predictor** |  |  |  |  |  |  |  |
| Physical activity | Yes (Ref) |  |  | — | — | — | — |
|  | No |  |  | 0.74 (0.53, 1.03) | 0.07 | 0.76 (0.53, 1.11) | 0.156 |
| PE class attendance | Yes (Ref) |  |  | — | — | — | — |
|  | No |  |  | 0.79 (0.59, 1.08) | 0.136 | 0.75 (0.53, 1.05) | 0.092 |
| Fruit consumption | Yes (Ref) |  |  | — | — | — | — |
|  | No |  |  | 0.85 (0.61, 1.19) | 0.344 | 0.87 (0.59, 1.27) | 0.459 |
| Vegetable consumption | Yes (Ref) |  |  | — | — | — | — |
|  | No |  |  | 1.29 (0.89, 1.86) | 0.182 | 1.16 (0.77, 1.76) | 0.473 |
| Soft drink consumption | No (Ref) |  |  | — | — | — | — |
|  | Yes |  |  | 0.94 (0.66, 1.35) | 0.745 | 0.99 (0.66, 1.48) | 0.954 |
| Fast food consumption | No (Ref) |  |  | — | — | — | — |
|  | Yes |  |  | 0.98 (0.71, 1.36) | 0.907 | 1.00 (0.70, 1.44) | 0.989 |
| Salty snack consumption | No (Ref) |  |  | — | — | — | — |
|  | Yes |  |  | 0.83 (0.60, 1.15) | 0.256 | 0.78 (0.54, 1.11) | 0.167 |
| Fatty food consumption | No (Ref) |  |  | — | — | — | — |
|  | Yes |  |  | 0.99 (0.72, 1.37) | 0.969 | 1.05 (0.73, 1.50) | 0.79 |
| Nutritional Knowledge | High (Ref) |  |  | — | — | — | — |
|  | Low |  |  | 1.09 (0.79, 1.50) | 0.611 | 1.17 (0.81, 1.69) | 0.4 |
| Diet quality | High(Ref) |  |  | — | — | — | — |
|  | Low |  |  | 0.98 (0.68, 1.41) | 0.923 | 1.26 (0.85, 1.86) | 0.258 |

*\*Statistically significant at p < 0.05. Model 1 is adjusted for demographic factors; Model 2 is adjusted for behavioral and dietary factors; and Model 3 is adjusted for both demographic and dietary factors.*

### Co-existence of double burden of malnutrition

In the fully adjusted model, low nutritional knowledge and low diet quality were significantly associated with the coexistence of the double burden of malnutrition. Adolescents with low nutritional knowledge had approximately two-fold higher odds of the double burden than those with high nutritional knowledge (AOR = 2.16, 95% CI: 0.99–4.71; p=0.041). Similarly, those with low diet quality had more than five-fold higher odds of experiencing the double burden (AOR = 5.27, 95% CI: 2.42–11.48; p<0.001). No other demographic, behavioral, or dietary factors were significantly associated with the double burden in the fully adjusted model.

**Table 4.** Multivariable binary logistic regression analyses for factors associated with the coexistence of the double burden of malnutrition among school-going adolescents in Kathmandu Valley, Nepal.

|  |  | **Model 1** |  | **Model 2** |  | **Model 3** |  |
|---|---|---|---|---|---|---|---|
| **Predictor** | **Category** | **Adjusted OR (95% CI)** | **p-value** | **Adjusted OR (95% CI)** | **p-value** | **Adjusted OR (95% CI)** | **p-value** |
| Sex | Male (Ref) | — | — |  |  | — | — |
|  | Female | 1.04 (0.51, 2.14) | 0.91 |  |  | 1.03 (0.48, 2.19) | 0.946 |
| Age category | 10–14 (Ref) | — | — |  |  | — | — |
|  | 15–19 | 0.57 (0.28, 1.18) | 0.131 |  |  | 0.48 (0.22, 1.05) | 0.068 |
| Caste | Advantage ethnic group (Ref) | — | — |  |  | — | — |
|  | Disadvantage ethnic group | 1.28 (0.56, 2.90) | 0.557 |  |  | 1.09 (0.46, 2.57) | 0.851 |
| Mother’s education | High (Ref) | — | — |  |  | — | — |
|  | Low | 1.70 (0.75, 3.87) | 0.206 |  |  | 1.37 (0.58, 3.24) | 0.475 |
| Father’s education | High (Ref) | — | — |  |  | — | — |
|  | Low | 1.56 (0.68, 3.57) | 0.29 |  |  | 1.36 (0.57, 3.24) | 0.493 |
| Mother’s occupation | Job/business (Ref) | — | — |  |  | — | — |
|  | Agriculture | 1.02 (0.46, 2.24) | 0.968 |  |  | 0.91 (0.40, 2.08) | 0.82 |
| Father’s occupation | Job/business (Ref) | — | — |  |  | — | — |
|  | Agriculture | 1.94 (0.88, 4.26) | 0.099 |  |  | 1.88 (0.81, 4.34) | 0.141 |
| **Predictor** |  |  |  |  |  |  |  |
| Physical activity | Yes (Ref) |  |  | — | — | — | — |
|  | No |  |  | 1.12 (0.56, 2.25) | 0.75 | 1.64 (0.74, 3.60) | 0.22 |
| PE class attendance | Yes (Ref) |  |  | — | — | — | — |
|  | No |  |  | 1.57 (0.76, 3.25) | 0.228 | 1.39 (0.62, 3.16) | 0.426 |
| Fruit consumption | Yes (Ref) |  |  | — | — | — | — |
|  | No |  |  | 1.35 (0.63, 2.90) | 0.438 | 1.58 (0.65, 3.81) | 0.31 |
| Vegetable consumption | Yes (Ref) |  |  | — | — | — | — |
|  | No |  |  | 1.85 (0.84, 4.04) | 0.125 | 1.28 (0.53, 3.11) | 0.582 |
| Soft drinks | No (Ref) |  |  | — | — | — | — |
|  | Yes |  |  | 1.76 (0.78, 3.96) | 0.171 | 2.04 (0.80, 5.18) | 0.133 |
| Fast food consumption | No (Ref) |  |  | — | — | — | — |
|  | Yes |  |  | 0.87 (0.40, 1.89) | 0.718 | 1.16 (0.48, 2.79) | 0.747 |
| Salty snack consumption | No (Ref) |  |  | — | — | — | — |
|  | Yes |  |  | 0.66 (0.30, 1.43) | 0.289 | 0.50 (0.20, 1.26) | 0.143 |
| Fatty food consumption | No (Ref) |  |  | — | — | — | — |
|  | Yes |  |  | 1.72 (0.81, 3.69) | 0.161 | 1.54 (0.64, 3.70) | 0.331 |
| Nutritional Knowledge | High (Ref) |  |  | — | — | — | — |
|  | Low |  |  | 2.17 (1.09, 4.31) | 0.027* | 2.16 (0.99, 4.71) | 0.041* |
| Diet quality | High (Ref) |  |  | — | — | — | — |
|  | Low |  |  | 4.24 (2.14, 8.40) | <0.001* | 5.27 (2.42, 11.48) | <0.001* |

*\*Statistically significant at p < 0.05. Model 1 is adjusted for demographic factors; Model 2 is adjusted for behavioral and dietary factors; and Model 3 is adjusted for both demographic and dietary factors. DBM was defined as the concurrent presence of stunting (HAZ < –2 SD) and overweight/obesity (BAZ > +1 SD).*

## Discussion

The present study provides evidence on the burden and correlates of malnutrition among urban school-going adolescents in the Kathmandu Valley of Nepal, highlighting the coexistence of undernutrition and overweight/obesity within the same population. Overall, 15.3% of adolescents were stunted, 5.5% were underweight, 16.9% were overweight/obese, and 3.0% experienced the coexistence of the DBM. This study explored the prevalence of DBM and its determinants among school-going adolescents within the Kathmandu Valley of Nepal and found a DBM prevalence of 3.0%, which is less than half of what has been observed in the secondary data analysis of the Nepal Demographic and Health Survey, 2016^13^ and study of Sunuwar et al.^44^ Although there are no known primary studies specifically assessing DBM among adolescents in Nepal, comparable findings have been observed in a large scale analysis of 57 LMICs adolescents in South Asian countries’ DBM where the prevalence of concurrent stunting and overweight-DBM was 2%^9^, which is tenfold of what was observed among Brazilian adolescents where it was only 0.2%^16^. This slightly higher prevalence could be attributed to dietary shifts, sedentary lifestyle and socioeconomic changes specific to the Kathmandu Valley, Nepal^45^.

A particularly important finding of this study was the strong association between age and stunting. Adolescents aged 15–19 years had approximately twice the odds of being stunted compared with those aged 10–14 years (AOR = 2.03, 95% CI: 1.35–3.06; p<0.001). This finding is consistent with evidence from South Asian adolescents showing a higher prevalence of stunting during late adolescence^46, 47^ and with findings from Ethiopia reporting greater odds of stunting among older adolescents^16^. The higher odds observed among older adolescents should, however, not necessarily be interpreted as indicating that older age itself causes stunting. Stunting reflects cumulative growth faltering and may therefore represent nutritional disadvantages experienced earlier in the life course. In addition, differences in pubertal timing and the increased nutritional requirements associated with adolescent growth may influence anthropometric status during this period. The finding consequently highlights the importance of addressing nutritional vulnerability before and throughout adolescence rather than viewing adolescent stunting solely as a problem emerging during late adolescence.

The strongest modifiable correlates of stunting in the fully adjusted model were observed to be diet quality, and nutritional knowledge. Adolescents with low diet quality had more than seven times higher odds of stunting than those with high diet quality (AOR = 7.27, 95% CI: 4.96–10.65; p<0.001), representing the strongest association identified in the stunting model. Similarly, adolescents with low nutritional knowledge had more than three times higher odds of stunting (AOR = 3.17, 95% CI: 2.17–4.66; p<0.001). These findings are consistent with evidence from Nepal and other settings indicating an association between inadequate nutritional knowledge, poorer dietary practices, and adolescent undernutrition^48 49^. Dietary quality is particularly important during adolescence because rapid growth increases requirements for energy, protein, and micronutrients. Diets characterized by limited diversity or inadequate nutritional quality may therefore be insufficient to support optimal linear growth. Previous evidence has similarly linked greater dietary diversity and improved diet quality with better nutritional outcomes among adolescents^50 51 52^.

The strong association between nutritional knowledge and stunting may reflect the potential role of knowledge in shaping food choices and dietary practices. Adolescents with greater nutrition knowledge may be more likely to recognize nutrient-dense foods and adopt healthier dietary practices^53^. However, knowledge alone is unlikely to determine nutritional status. The ability to translate knowledge into dietary behavior may depend on household food availability and affordability, parental practices, school food environments, and broader socioeconomic conditions. Thus, the observed association supports an integrated approach in which nutrition education is combined with improvements in the food environment and access to diverse and nutritious foods.

Consumption of salty snacks was also associated with higher odds of stunting in the fully adjusted model (AOR = 1.53, 95% CI: 1.00–2.36; p=0.031). Although the magnitude of this association was substantially smaller than that observed for diet quality and nutritional knowledge, it may indicate the influence of broader dietary patterns among adolescents. Frequent consumption of energy-dense, nutrient-poor snack foods may displace nutrient-dense foods within the diet and may therefore be associated with poorer overall dietary adequacy. This finding also reinforces the importance of considering the adolescent food environment, particularly the availability and consumption of commercially prepared snack foods around schools. However, because the present study did not establish temporality, the observed association should be interpreted as correlational rather than causal.

An inverse association was also observed between fatty-food consumption and stunting (AOR = 0.65, 95% CI: 0.42–1.00; p=0.049). This finding should be interpreted cautiously. The category of “fatty foods” may include foods with substantially different nutritional profiles, ranging from nutrient-rich sources of dietary fat to fried or processed foods. Consequently, the observed inverse association may reflect differences in the types of foods included in this category, overall dietary patterns, or residual confounding rather than a protective effect of fatty-food consumption itself. A study done among school age children in North Lebanon found the association of double burden of malnutrition with unhealthy dietary behavior^54^. Further research using more detailed dietary assessment would be required to clarify this relationship.

In contrast to the strong associations observed for stunting, the fully adjusted analysis of body-weight status identified considerably fewer independent correlates. The prevalence of overweight/obesity was 16.9%, demonstrating that excess weight is also an important nutritional concern among adolescents in the Kathmandu Valley. The observed age-related pattern is broadly consistent with evidence from China and South Asian populations indicating differences in overweight/obesity prevalence across adolescent age groups^9 55^. Such differences may reflect changes in growth, physical activity, dietary behavior, and body composition during adolescence. Nevertheless, interpretation of the regression finding should remain closely aligned with the exact coding of the dependent variable and the final adjusted model. Importantly, the relatively high prevalence of overweight/obesity alongside stunting in the same population illustrates the coexistence of different forms of nutritional vulnerability during adolescence.

The finding of a 3.0% prevalence of DBM is particularly important because it demonstrates that undernutrition and excess weight are not necessarily separate public health problems within this population. Rather, they may coexist within the same nutritional and social environment. The fully adjusted model identified low diet quality as the strongest correlate of DBM. Adolescents with low diet quality had more than five times higher odds of experiencing the DBM compared with those with high diet quality (AOR = 5.27, 95% CI: 2.42–11.48; p<0.001). Low nutritional knowledge was also associated with approximately two-fold higher odds of DBM (AOR = 2.16, 95% CI: 0.99–4.71; p=0.041), consistent with the study done among adolescents in Zimbabwe ^56^. Together, these findings suggest that the double burden may be linked to common underlying dietary and knowledge-related factors rather than representing two entirely independent nutritional problems.

The strong association between poor diet quality and DBM is particularly relevant in the context of the nutrition transition. A diet may provide sufficient or excessive energy while remaining inadequate in essential nutrients, particularly when consumption of energy-dense foods occurs alongside inadequate dietary diversity. Such dietary patterns may create conditions in which undernutrition and excess weight coexist. The present findings therefore support a broader conceptualization of adolescent nutrition in which the quality and composition of the diet are considered alongside total energy intake. Further, the association between low nutritional knowledge and DBM suggests that adolescents’ understanding of healthy nutrition may be relevant to the emerging double burden. However, as with stunting, nutrition knowledge should not be considered an isolated determinant. Adolescents make food choices within environments shaped by household resources, food prices, availability, marketing, school food environments, and prevailing social norms. An integrated strategy combining nutrition education with supportive school and household food environments may therefore be more appropriate for addressing the multiple forms of malnutrition observed in this population.

Several limitations should be considered when interpreting these findings. First, the cross-sectional design limits the ability to establish temporality or causal relationships between dietary, behavioral, and socioeconomic factors and nutritional outcomes. In particular, reverse causality cannot be excluded, as existing nutritional status may itself influence dietary practices and behaviors. Second, dietary and behavioral information was collected using interviewer-administered questionnaires and may therefore be subject to recall and social desirability bias. Third, the study was restricted to urban, school-going adolescents in the Kathmandu Valley, limiting the generalizability of the findings to adolescents who are out of school and those living in rural or geographically different areas of Nepal. In addition, the use of fixed anthropometric classifications across a heterogeneous adolescent age range may not fully capture individual variation in pubertal growth and may contribute to some degree of nutritional-status misclassification.

Despite these limitations, the study has several strengths. It included a relatively large sample and used a multistage cluster sampling approach with probability proportional to size, which strengthens the representativeness of the sampled school-going adolescent population within the study setting. Standardized anthropometric measurements based on WHO growth references also enhance comparability with other studies. Furthermore, the inclusion of demographic, behavioral, dietary, and nutritional-knowledge variables allowed examination of multiple potential correlates of adolescent malnutrition within a common analytical framework. Multivariable logistic regression enabled adjustment for several measured covariates, thereby reducing the potential influence of measured confounding.

## Conclusion

Overall, this study highlights the complexity of adolescent malnutrition in urban Nepal. While stunting and overweight/obesity remain important and seemingly contrasting nutritional concerns, the coexistence of these conditions demonstrates the emergence of a broader double burden. The particularly strong associations of low diet quality and low nutritional knowledge with stunting and DBM suggest that improving the quality of adolescents’ diets, together with creating supportive environments that enable healthier food choices, may be central to addressing this challenge. Longitudinal and nationally representative studies are needed to further clarify the pathways linking dietary practices, socioeconomic conditions, pubertal development, and nutritional outcomes across adolescence and to evaluate integrated interventions capable of addressing multiple forms of malnutrition simultaneously.

## Author’s contribution

Dev Ram Sunuwar (DRSu), Ashmita Kiwanayo (AK), Subekshya Karki (SK), Asmita Shrestha (AS), Melisha Shrestha (MS), Dirghayu KC (DKC), Sudip Sunuwar (SS), Devendra Raj Singh (DRSi), Kshitij Karki (KK), Narendra Kumar Chaudhary (NKC). Conceived and designed the study: **DRSu**, **AK, SK, AS, MS, DRSi, KK, NKC, SS, and DKC**. Tools translation to the Nepali language and back translation: **AK, SK, MS, and AS**. Data collection: **AK, SK, MS, and AS**. Analyzed the data: **AK, SK, AS, MS, SS, and DRSu.** Writing and editing the manuscript: **DKC, DRSi, and DRSu**. Critical revision of the manuscript: **DKC, DRSi, NKC, KK, and DRSu**. All authors have read and approved the manuscript. DKC accepts full responsibility for the finished work, had access to the data, and controlled the decision to publish. DKC is the guarantor of this work.

## Acknowledgement

The authors would like to acknowledge the participants’ valuable participation in the study. Also, we would like to thank anyone who directly or indirectly contributed to and supported our study, including teachers and parents of the students and students at the respective schools, for their facilitation and cooperation in undertaking this research.

## Patients and Public involvement

Neither patients nor the public were involved in the design, conduct, reporting, or dissemination of this research.

## Patient consent for publication

Not applicable.

## Data availability statement

Data are available on reasonable request.

## Ethical approval

Ethical approval for this study was sought from the Nepal Health Research Council (Protocol registration number: 144\2022p). Written permission letters from each participating school to conduct the study were also secured prior to stating the study. Before data collection, written informed consent from guardians, and written assent from participants were secured.

## Supplementary material

Supplementary files containing additional data and materials relevant to the study have been uploaded alongside the manuscript. These files provide further details that support the findings described in the manuscript as well as enhance the transparency and reproducibility of the research.

## Abbreviations

DBM: Double Burden of Malnutrition
LMICs: Low- and Middle-Income Countries
WHO: World Health Organization
UNICEF: United Nations Children’s Fund
GSHS: Global School Based Student Health Survey
NMSS: Nepal National Micronutrient Survey
PPS: Probability Proportional to Size
IPAQ: International Physical Activity Questionnaire
pHDI: pro-Healthy Diet Index
nHDI: non-Healthy Diet Index
BMI: Body Mass Index
NHRC: Nepal Health Research Council
NDHS: Nepal Demographic and Health Survey

